# Revisiting biological sex as a risk factor for COVID-19: a fact or mirage of numbers?

**DOI:** 10.1101/2022.07.02.22276577

**Authors:** Samer Singh

## Abstract

Biological sex is considered a risk factor for COVID-19. The prevailing view supposes males are about two-fold more impacted than females based on early-stage studies. The observed higher male deaths in COVID-19 are purportedly a result of biological differences that make males more vulnerable to adverse outcomes in infectious diseases. Research and policy paradigms seem to follow a similar line of thought to mitigate COVID-19 impact on populations. The analysis of sex-disaggregated data could help us evaluate the veracity of assertions for a preferred evidence-guided response. The analysis of the sex-disaggregated data available for the top 70 countries contributing about 80% of total deaths (as of 15 September 2021; on average two waves of infections experienced) indicates average Case Sex (Male: Female) ratio (CSR) of 1.09±0.35 (marginally more male cases) and Death Sex ratio (DSR) of 1.48± 0.47. Consideration of only laboratory-confirmed cases indicates the mortality sex ratio (MSR) in COVID-19 (MSR-COVID) to be 1.37±0.30. The prevailing MSR for the same countries was 1.758±0.409. The relative change in the mortality rate for males as compared to females in COVID-19 (ratio: MSR-COVID/prevailing MSR-PP) was 0.818±0.261 much lower than anticipated (2 or higher). Overall, over three-fold more countries (51/70) experienced a higher rate of female mortality than male mortality (15/70). Together, it suggests a more disproportionately severe impact of COVID-19 on females than on males, contrary to the prevailing view. Identification and analysis of country-specific factors contributing to differential impact on sexes, whether biological or environmental, seem warranted.

## INTRODUCTION

The global impact of COVID-19 on sexes has been differential in populations. Early small-scale studies had indicated more SARS-CoV-2 infections caused reported deaths among males as compared to females [1-6]. Females are generally estimated to be about two-fold protected from COVID-19 adverse outcomes. Differential incidence and outcomes in populations had been purported to be resulting from underlying biological differences, e.g., immune system, ACE2 receptor expression, polymorphism, etc. [7-11]. The notion of males being at risk still seems to be driving our ongoing research directions, focusing on the biology and immune system of females, and possibly the policies for COVID-19 control and impact minimization as well [12-17], without reanalyzing the existing data and taking into consideration the fact that higher death rate in males is globally pervasive. The adult mortality sex (Male: Female) ratio (MSR) differences among countries remain quite large, ranging from about 1 to 3 with region-specific trends [18,19]. The differential higher rate of male deaths is attributed more to the behavioral patterns and associated prevailing environmental risk factors of males specific to countries than the biological differences *per se [20-24]*. As huge differences in the sex-specific mortality rates exist across countries due to their inherent contributory factors, the differences observed in the sex-specific mortality rates in a few small study populations of COVID-19 alone cannot justify the depiction of one sex being inherently more protected than the other due to biological differences as it could be inherently misleading. The comparison of MSR observed in COVID-19 (MSR-COVID) with prevailing pre-pandemic MSR (MSR-PP) for countries could be better placed to identify the potential role of biological sex on the COVID-19 impact if any.

As of December 10th, 2021, a total of 271.26 million COVID-19 cases and 5.34 million associated deaths have been reported [25]. However, the availability of age and sex-disaggregated data remains poor. The sex-disaggregated data of COVID-19 impact on populations collated by Global Health 5050 [26]–a consortium for equity in health that tracks global sex-disaggregated country-specific data on COVID-19, in conjunction with the data on pre-pandemic adult MSR from Global Health Observatory, WHO 2018 [18], could be a credible source of such data for reassessing the prevailing view on the role of biological sex on COVID-19 impact on populations (Table 1).

**Table 1.**
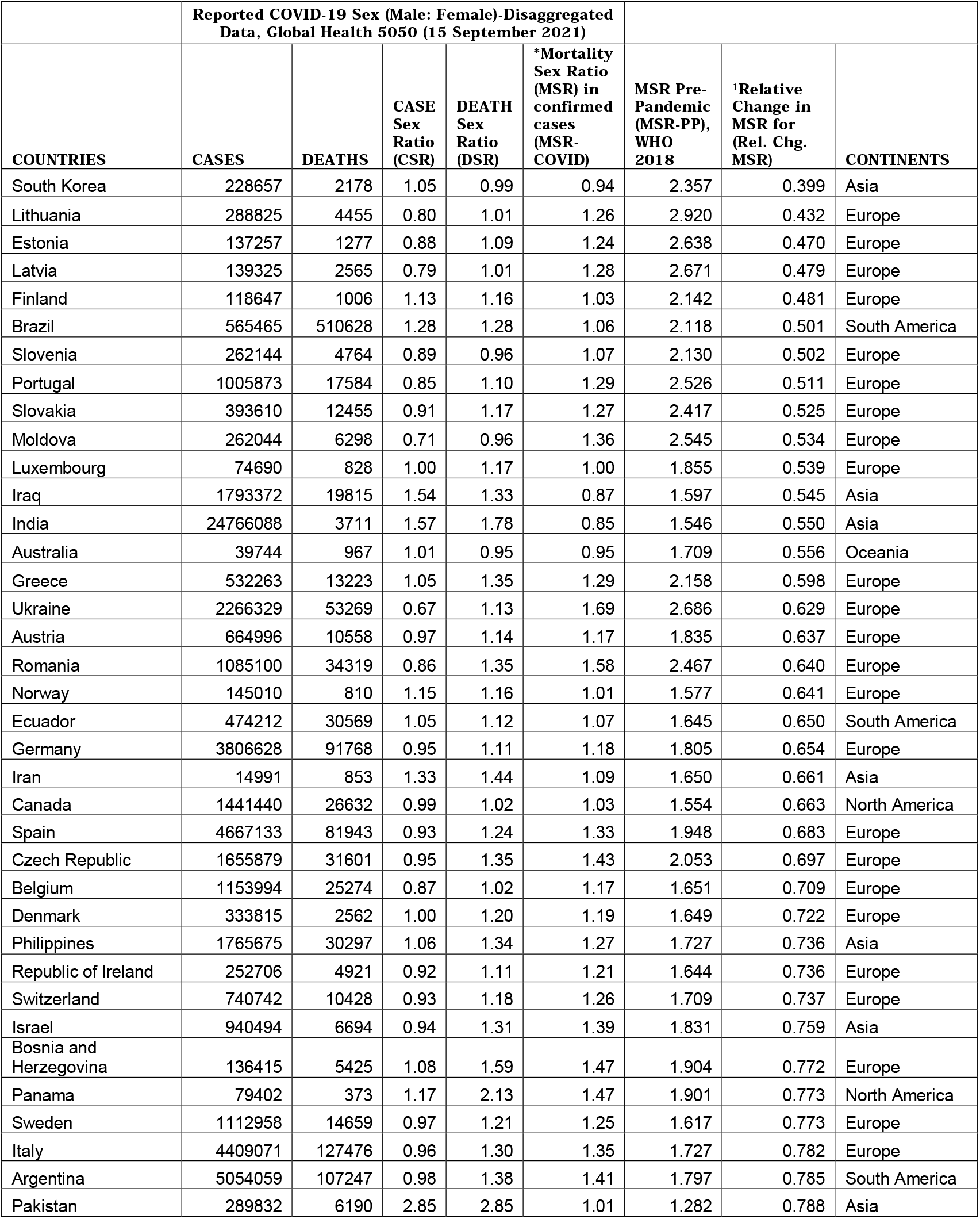
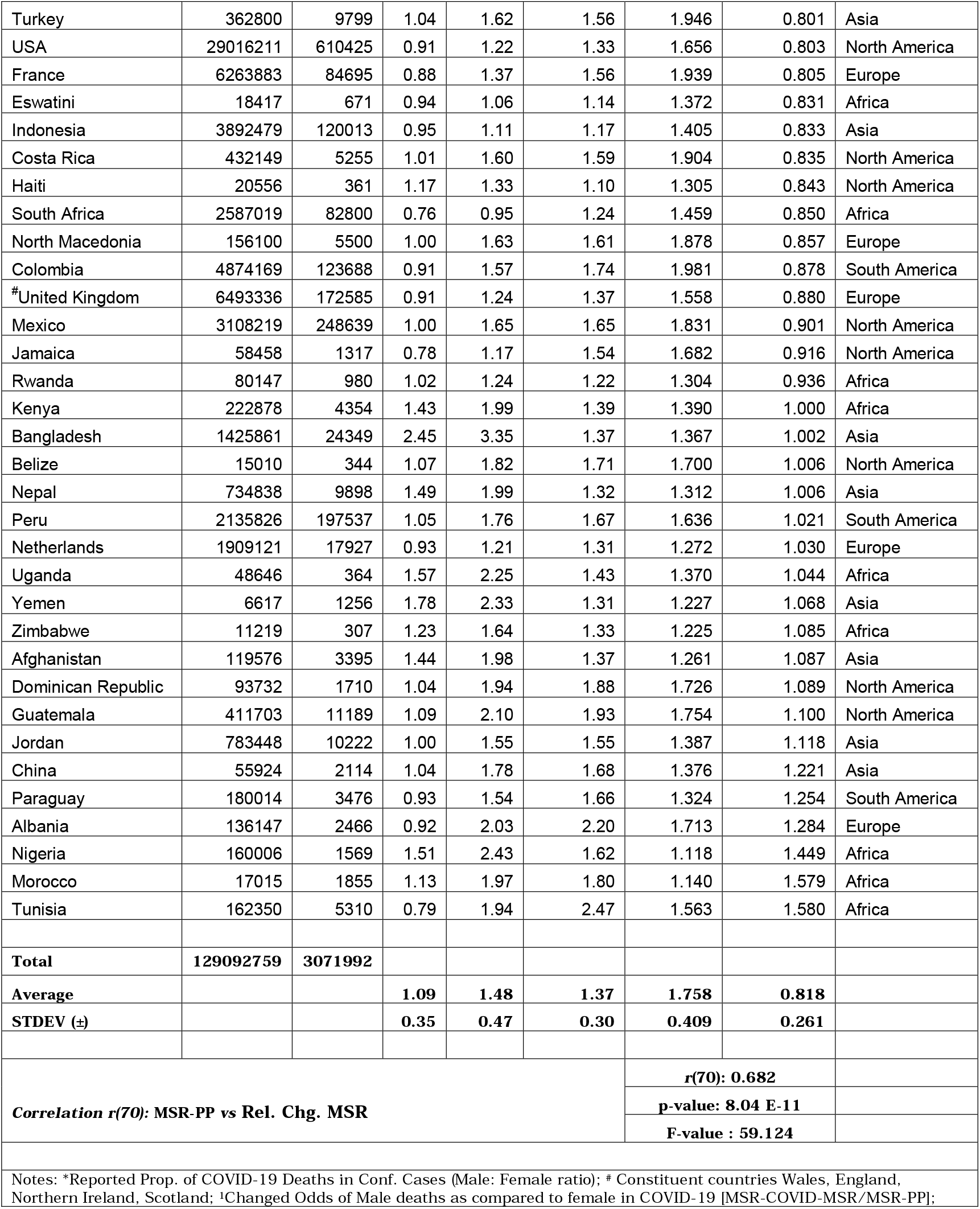
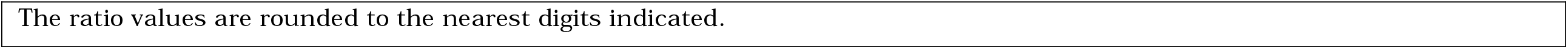
COVID-19 sex-disaggregated data, adult mortality sex ratios, and covariation analysis. The COVID-19 data and adult mortality sex ratio from refs [26] and [18] respectively.

## RESULTS AND DISCUSSION

A cursory look at country-wise COVID-19 incidence and death data along with the observed raw Cases Sex ratio (CSR), Deaths Sex Ratio (DSR), and the Mortality Sex ratio in laboratory-confirmed cases (MSR-COVID) of top 70 countries (from [26] as on 15 September 2021, Table 1) that are contributing about 80% of global deaths, indicates countries in Europe and North America are reporting more female cases while that in Asia and Africa reporting male cases. When we look at reported COVID-19 incidence/cases sex ratio (CSR) across countries, it is found to vary widely with average value of 1.09 ± 0.35 [range: 2.85 (Pakistan) - 0.67(Ukraine)] (Table 1; Fig. 1). Though the number may leave us thinking of more male cases in general, the distribution of countries with CSR values below and above 1 seemed even. Out of 70 countries, the countries reporting CSR values>1.00, 1.00, and <1.00 were 32, 5, and 33 countries, respectively. The distribution of reported incidences of COVID-19 globally does not seem to support any notion of one sex being particularly more prone than the other to contracting SARS-CoV-2 despite the data displaying country-specific propensities. The reported incidence variations between sexes among countries seem to be predominantly reflective of countries prevailing sex (Male: Female) ratio (*e*.*g*., more females in countries of Europe and North America while males in the countries of Asia and Africa). Additionally, natural variation arising from potential occupational differences/hazards, policies to report or test individuals, health-seeking behavior, *etc*. from country to country could be also contributing to some small differences.

**Figure 1.**
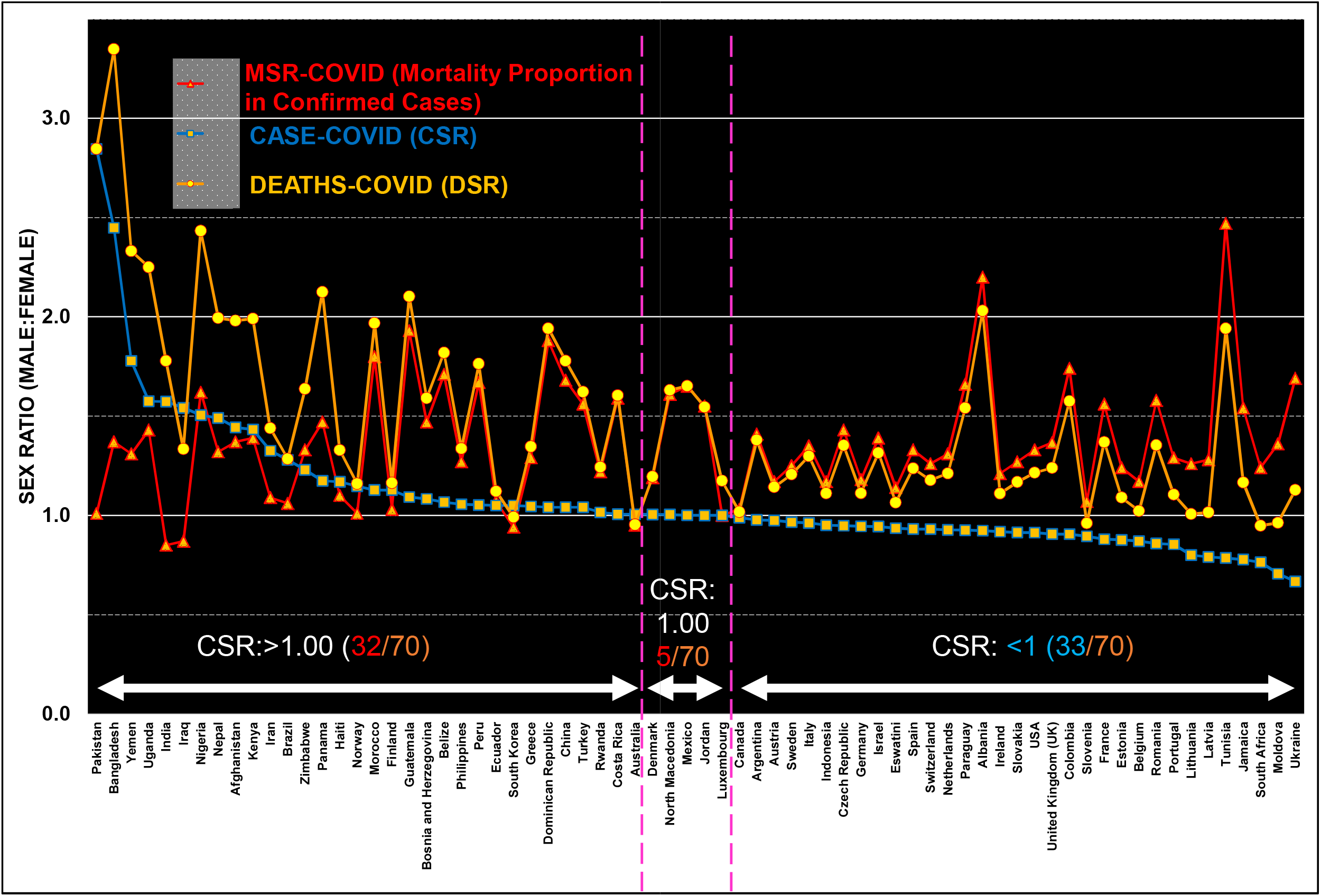
Sex (Male: Female) Ratio in reported COVID-19 incidences/cases, deaths, and mortality proportion in laboratory-confirmed COVID-19 cases. Data of the top 70 countries accounting for about 80% of deaths and reporting sex-disaggregated data (as of 15 September 2021) is presented.

The sex ratio of COVID-19 deaths reported or Death Sex Ratio (DSR) reported by the same countries from the available sex-disaggregated deaths paints a much grimmer picture (Table 1). The reported COVID-19 deaths sex ratio (DSR) among countries displayed more extremes with an average value of 1.48±0.47 [range: 3.35 (Bangladesh) - 0.95 (South Africa)] and most nations reporting DSR >1.00, *i*.*e*., more males dying than females. As different countries have differential infection rates for sexes, the ratio of mortality proportion in the confirmed male and female COVID-19 cases or mortality sex ratio in COVID-19 (MSR-COVID) would be appropriate to be compared across countries to assess differential sex-dependent mortality among countries if any. The comparison of reported/calculated MSR-COVID among countries presents a less extreme differential mortality among sexes across countries. The MSR-COVID for the countries averaged 1.37±0.30 [range: 2.47 (Tunisia) to - 0.85 (India)] with an overwhelming number of countries (66/70) showing a ratio of > 1.00, indicating disproportionate deaths of males in COVID-19. Interestingly, a few countries with MSR-COVID <1.00 (*i*.*e*., higher rate of mortality among female COVID-19 cases) were India (0.85), Iraq (0.87), Australia (0.95), and South Korea (0.94). This group of countries seemed to be an aberration to what is being observed all over the world. Some other countries that displayed near-neutral covid adverse impact on sexes (*i*.*e*., MSR-COVID: 1.00 - 1.05) were Luxembourg (1.00), Pakistan (1.01), Norway (1.01), Canada 1.03, Finland 1.03. Here readers may be again reminded of the existence of a huge difference in mortality sex ratio (MSR) globally (range: about 1 to 3) among countries with significant region-specific variations ranging from an average of 1.2 for sub-Saharan African countries to an average of 2.1 for North American and European nations [18,19]. The huge difference in the MSR for countries is attributed to existing underlying risky behaviors, whereas biological differences are supposed to have constant but very small to negligible contributions [20-22]. Since the biology of females is asserted to be protecting females or alternatively putting the males at an additional risk of dying from COVID-19, it would be reasonable to compare the COVID-19 specific MSR (MSR-COVID) with the pre-pandemic levels of MSR for countries (MSR-PP) to reach a conclusion.

The estimated MSR-PP for the countries as per WHO, 2018 [18] are presented in Table 1. In case biological differences attributed to the female sex were making a positive contribution to female survival, *i*.*e*., less impacted by COVID-19, the MSR-COVID would be expected to be higher than the prevailing MSR-PP for countries. The average MSR-PP for countries included in the current analysis was found to be 1.758±0.41 [range: 2.920 (Lithuania) - 1.118(Nigeria)] well above the average MSR-COVID of 1.37±0.30 for countries (Table 1). Both the MSR-PP and MSR-COVID for countries are graphically depicted in Fig 2. Contrary to what would be expected from the prevailing notion, for the significant majority of the countries, i.e., 51/70, the MSR-COVID remained lower than MSR-PP, signifying the overwhelming majority of countries were witnessing a higher rate of female deaths than expected, while a small minority 19/70 countries only displayed male mortality equal to the prevailing levels or higher. To give an idea of the relative change in the mortality rates for males in comparison to females during COVID-19 in relation to the prevailing mortality rate sex ratio, the ‘Rel. Chg. MSR’ (ratio: MSR-COVID/MSR-PP) for countries is also indicated in Fig. 2. A value greater than 1.00 would indicate a higher risk of male deaths, while a value smaller than 1.00 should indicate more risk of female deaths than expected from the prevailing MSR for countries, even when any protective contribution from the differences in the biology of the sexes is not taken into account. Out of 70 countries, 51, 4, and 15 had the ratio of <1.00, 1.00-1.01, and >1.01, respectively. The majority of countries 55/70 displayed reduced or no significant change in male mortality. The ‘Rel. Chg. MSR’ averaged 0.818±0.261 for countries indicating the overall differential lower male death rates in COVID-19 than expected among countries had the pre-pandemic levels of expected biological advantage remained with females as compared to males. The relative change in MSR for COVID-19 negatively covaried with the countries prevailing MSR-PP [*r*(70): 0.682, p-value: 8.06 E-11, F-value 59.112; Table 1). Had the biology been protecting females from COVID-19, we would not be seeing globally almost equal distribution of countries with regard to predominant male and female infections whereas a higher rate of female deaths in over 2.5-fold more countries than those displaying male deaths at pre-pandemic level or higher from the expected of prevailing MSR. These differentials predominantly higher rate of female deaths combined with even distribution of infection rates among sexes question the very notion of biology protecting females from SARS-CoV-2 infections and mortality.

**Figure 2.**
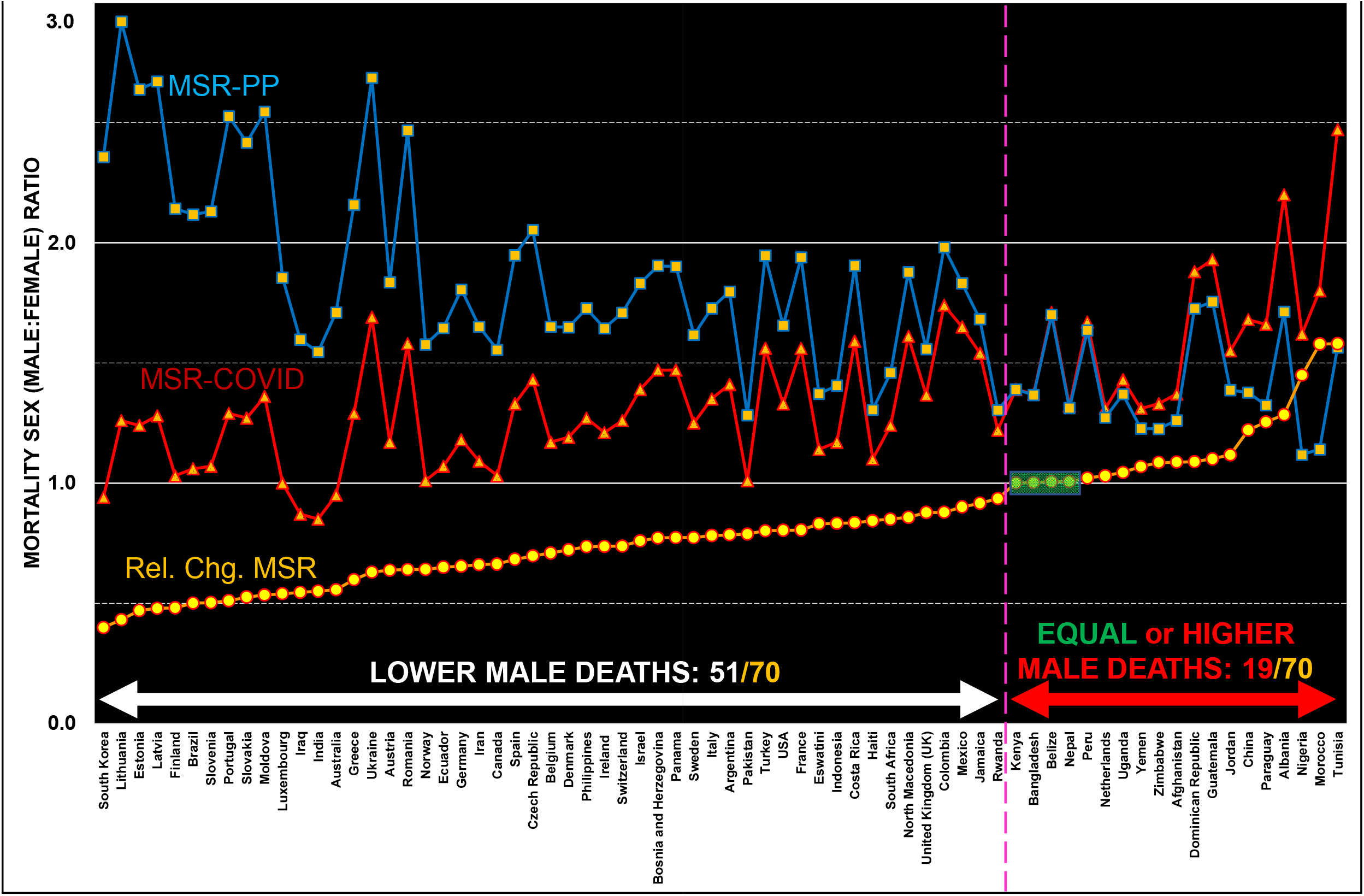
Relative change in the mortality sex ratio of countries for COVID-19 to that of preexisting (Rel. Chg. MSR: MSR-COVID/MSR-PP) indicates lower male deaths in the majority of countries. A majority of countries (51/70) reported lowered rate of male death rates while a small minority of countries (19/70) displayed no change (4; green boxed) to a higher rate (15) of male mortality in COVID-19, mostly from Africa and Asia. A vertical magenta line separates countries displaying lower and equal or higher rate of male mortality (deaths) in COVID-19 than the prevailing rates.

Countries with higher prevailing pre-pandemic MSR (MSR-PP) seem to predominate the group of countries that display relative change in MSR during COVID-19 to smaller values, *i*.*e*., less male mortality rates while more female mortality rates than expected (Fig. 2). Expectedly, a plot of countries with decreasing order of MSR-PP indicates an inverse correlation between relative mortality sex ratio indicator for COVID-19 (Rel. Chg. MSR) and MSR-PP, *i*.*e*, higher MSR-PP of countries associated with lower male mortality in COVID-19 (Fig. 3). Among the countries falling in the prevailing MSR-PP brackets of <1.40, 1.40- <1.79, and >1.79, only 10/17, 5/25 and 0/28 countries displayed higher male mortality in confirmed COVID-19 cases, indicating the vulnerability of males to COVID-19 deaths progressively predominate in countries with lower MSR-PP while more female vulnerability to predominate countries of higher MSR-PPs. Interestingly, the countries with MSR-PP of 1.79 that exclusively display more female vulnerability, mostly belong to Europe and North America with higher Human developmental index and medical facilities [27].

**Figure 3.**
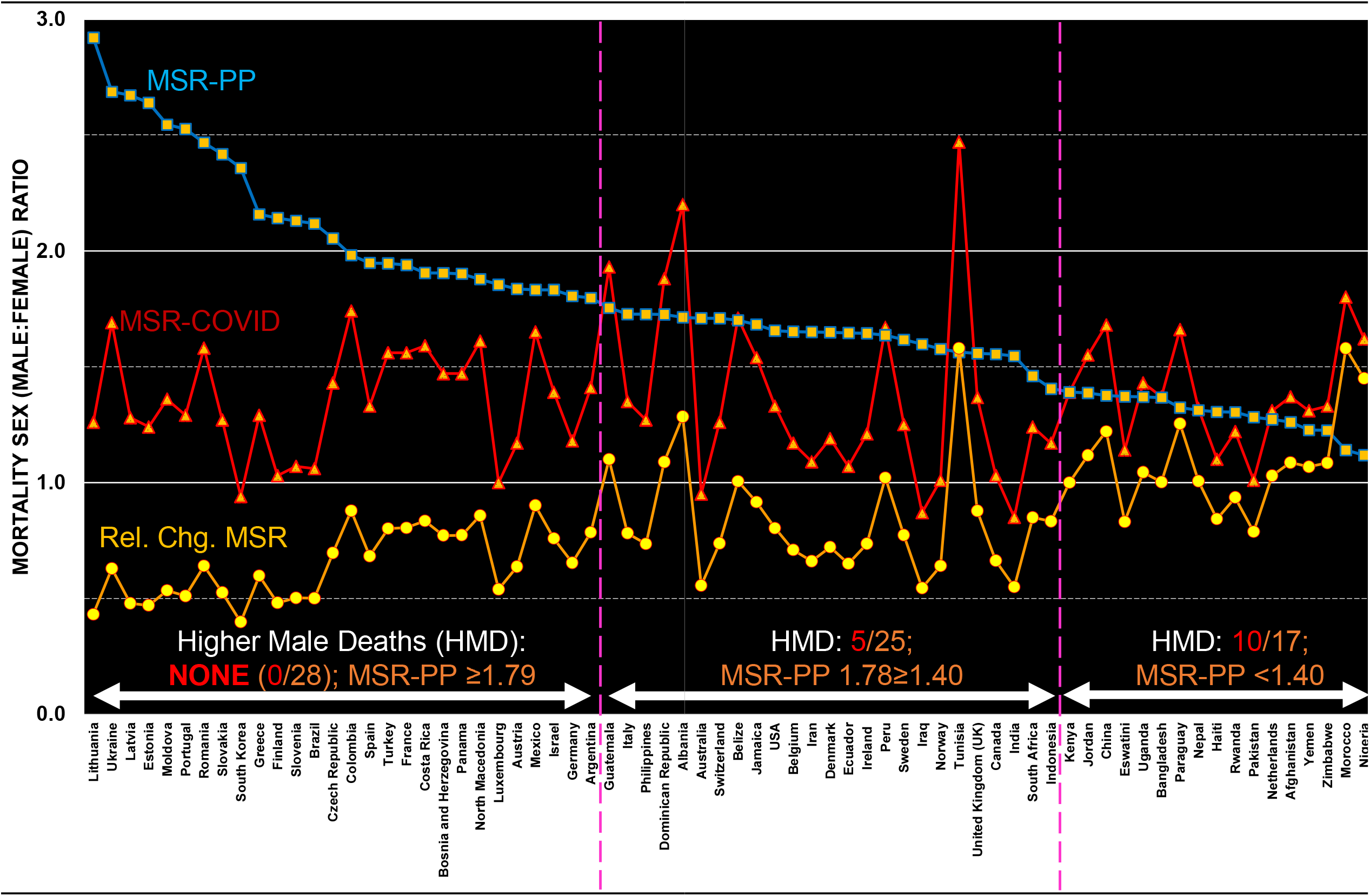
Variation of relative change in mortality sex ratio across countries in COVID-19 (Rel. Chg. MSR) with prevailing pre-pandemic MSR (MSR-PP). Out of a total of 70 countries, 28 falling in the MSR-PP brackets of >1.79 exclusively displayed higher female death rates, whereas just 5/24 countries falling in the MSR-PP bracket of 1.78 to ≥1.4 and 10/17 countries in MSR-PP bracket of < 1.4 displayed higher male death rate. Countries in different brackets of MSR-PPs demarcated by vertical magenta lines.

The observations presented have limitations in terms of presenting a complete global picture as the sex-disaggregated data reporting has not been uniform among countries. Currently, the data is available for only a subset of total cases for the majority of the reporting nations. Several countries are still not reporting sex-disaggregated data, while many that do so still lag in providing complete COVID-19 data disaggregated by sex and age in a reasonable timeframe [26; 2-6 months delay is common].

Combined together, the adverse outcomes in confirmed COVID-19 cases do not seem to support biological differences playing a major role in the overall impact of COVID-19 on population constituents. Could it be the prevalence of differential risk-factors/ protective-variables [28-31] or policy implementations [27] that is causing disproportionate female deaths? A more thorough assessment of the role of biological differences in COVID-19 impact on populations could be made possible by countries through comparable reporting of the sex-disaggregated data with regard to potential variables purported to be playing a role, *e*.*g*., age, morbidities, time to death, complications, medications, medical support available, *etc*. [26,27]. It would be interesting to know how these mortality sex ratios among countries would have varied with various proposed explanatory and protective variables [28-31] including widely adopted inappropriate nutrient supplementations and medications [32-37].

## CONCLUSIONS

The observations put forth starkly question the very validity of the notion of biological sex protecting females from SARS-CoV-2 infection and adverse outcomes or putting males at risk. Even if it is playing a role as expected, the observed differences cannot be reconciled by biology alone. Potentially, a major reconsideration of ideas driving our research efforts and policy directives to help mitigate the effect of COVID-19 on different sexes may need to be explored in a country-specific manner. It is worth noting that the adult MSR used in the current analysis may be grossly underestimating the negative impact on females, as the MSR generally increases steeply with age for the most affected age group of the population, *i*.*e*., > 65 years [38]. Dedicated studies need to be undertaken to identify country-specific factors responsible for differential responses — disproportionately affecting one sex or the other. A proactive approach from countries on the generation and sharing of a more detailed and complete sex-disaggregated data set would be warranted to fill the data gap required for evaluation and analysis of measures in place for evidence-based guidance of research directions and mitigation policies. It could help a more thorough analysis with country-specific applicable risk factors or potential protective variables for identifying country-specific risk factors that could be impacting one sex more than the other.

## MATERIALS & METHODS

The COVID-19 cases and deaths data are from Worldometers [25]. The country-specific sex-disaggregated data of COVID-19 is sourced from Global Health 5050 [26]. The mortality rate sex ratio for countries data is from Global Health Observatory, WHO 2018 [18]. Cases Sex Ratio (CSR), Death Sex Ratio (DSR), Mortality Sex Ratio [both in confirmed COVID-19 cases (MSR-COVID-19) and the prevailing pre-pandemic (MSR-PP)] are the calculated ratio of observed male to female in reported COVID-19 cases, reported COVID-19 deaths, and reported mortality rates, respectively. General mathematical and statistical calculations on data, *e*.*g*., sum, average, STDEV, ratio, correlation analysis, were performed in Excel as done previously [29,31,39].

## Data Availability

The sex-disaggregated data on COVID-19 publically available at https://globalhealth5050.org/covid19/
Other data produced in the present work are contained in the manuscript.

## Acknowledgments

The stimulating discussions on the subject with Prof. Rakesh K. Singh, Department of Biochemistry, Institute of Science, Banaras Hindu University, Varanasi-221005, India are acknowledged. The IoE seed grant support from Banaras Hindu University to SS is acknowledged.

## Funding

No funding was received for the work presented here.

## Competing interests

No competing interests to disclose.

## Data and materials availability

All data are available in the main text and indicated sources.

## Notes

### Competing Interest Statement

The authors have declared no competing interest.

### Funding Statement

This study did not receive any funding.

### Author Declarations

The study used sex-disaggregated data on COVID-19 publically available at https://globalhealth5050.org/covid19/

